# Phase Shift Between Age-Specific COVID-19 Incidence Curves Points to a Potential Epidemic Driver Function of Kids and Juveniles in Germany

**DOI:** 10.1101/2021.11.29.21267004

**Authors:** Hans H. Diebner

**Affiliations:** Dept. of Medical Informatics, Biometry and Epidemiology, Ruhr-Universität Bochum, 44780 Bochum, Germany

**Keywords:** COVID-19, SARS-CoV-2, incidence cross-correlation, phase shift

## Abstract

Mutual phase shifts between three German COVID-19 incidence curves corresponding to the age classes of children, juveniles and adults, respectively, are calculated by means of delay-cross-correlations. At the country level, a phase shift of −5 weeks during the first half of the epidemic between the incidence curves corresponding to the juvenile age class and the curve corresponding to the adult class is observed. The children’s incidence curve is shifted by −3 weeks with respect to the adults’ curve. On the regional level of the 411 German districts (Landkreise) the distributions of observed time lags are inclined towards negative values. Regarding the incidence time series of the juvenile sub-population, 20% of the German districts exhibit negative phase shifts and only 3% show positive shifts versus the incidence curves of the adult sub-population. Similarly for the children with 6% positive shifts. Thus, children’s and juveniles’ epidemic activity is ahead of the adults’ activity. The correlation coefficients of shifted curves are large (> 0.9 for juveniles versus adults on the country level) which indicates that aside from the phase shift the sub-populations follow a similar epidemic dynamics. Negative phase shifts of the children’s incidence curves during the first and second epidemic waves are predictors for high incidences during the current fourth wave with respect to the corresponding districts.

## 1 Introduction

The SARS-CoV-2 epidemic reached Germany in February 2020 accompanied by a continuous controversy regarding the legitimization of containment measures. In particular, school closure [1], mask wearing of children as well as regular testing in schools are extremely controversially debated [2, 3]. Based on arguments that children do not substantially contribute to drive the epidemic, a short period of online education has been quickly suspended. After the establishment of COVID-19 rapid antigen tests, it has been argued that regular testing in schools may reduce the infection risk since positive test results allow for immediate interventions [2]. However, an effective control is antagonized by the current (17 Nov 2021) epidemic reproduction speed. Given the low sensitivity of rapid SARS-CoV-2 antigen tests [4], a large reproduction number in schools entails an unmanageable high frequency of testing in order to efficiently control the epidemic.

Of note, there is increasing evidence taken from studies into seroprevalence that the actual SARS-CoV-2 incidence within the population of children and juveniles is higher than previously expected [5, 6, 7, 8], which may be a further reason for the rapid spread of infections. However, as shown in the sequel, already on the basis of registered incidence time courses it can be shown that the epidemic activity of the young sub-population is considerably ahead of the adults’ activity. We refrain from an in-depth interpretation since this would imply access to precise data on local containment measures in the past which is challenging to obtain if not impossible. However, the information on the mere fact that the children’s epidemic activity pattern precedes the adults’ dynamics is of great value for the design of future public health policies.

## 2 Materials and Methods

Age-specific weekly sampled COVID-19 7-day incidence (per 100,000 persons) time series up to the final observation/retrieval time *t*_*f*_ = 17 Nov 2021 are provided by the Robert Koch-Institute on a local (411 districts) level [9]. The following three age classes are used: [0-14], [15-19] and [20+] (in life years). The three incidence time series *I*_*k*_(*t*), *I*_*j*_(*t*) and *I*_*a*_(*t*) (*k* = kids, *j* = juveniles, *a* = adults) are analyzed by means of mutual delayed cross-correlations. Thus, the phase shift *δ* which maximizes the correlation between two incidence curves *I*_*x*_(*t* + *δ*) and *I*_*y*_(*t*) is estimated.

The time steps are given in weeks from the first week of 2020 onward.

Cross-correlations are calculated on the country level as well as for all 411 districts using the ccf-function of statistics programming language R version 4.1.2 ([10]). The estimated phase shifts are then used to predict the current epidemic activity (incidence) by means of a linear regression model (lm-function of statistics programming language R).

## 3 Results

### 3.1 Cross-Correlations on the Global German-wide Level for the First Two Waves

The three global (German-wide) incidence time series for children, juveniles and adults, respectively, are depicted in Figure 1. Phase shifts between the curves are apparent on visual inspection. Precise estimates of the three mutual cross-correlation functions for the first two waves only (*t <* 60 weeks, see the vertical line in Figure 1 that marks the separation of the intervals) are shown in Figure 2. A phase shift of *δ* = −5 weeks maximizes the correlation coefficient of 0.913 for *I*_*j*_(*t* + *δ*) and *I*_*a*_(*t*). Analogously, we observe a phase shift of −3 weeks between *I*_*k*_ and *I*_*a*_ (correlation coefficient 0.932), and a phase of 2 weeks between *I*_*k*_ and *I*_*j*_ (correlation coefficient 0.982). Thus, both juveniles as well as children unfold an epidemic activity which is ahead of the adults’ activity.

**Figure 1:**
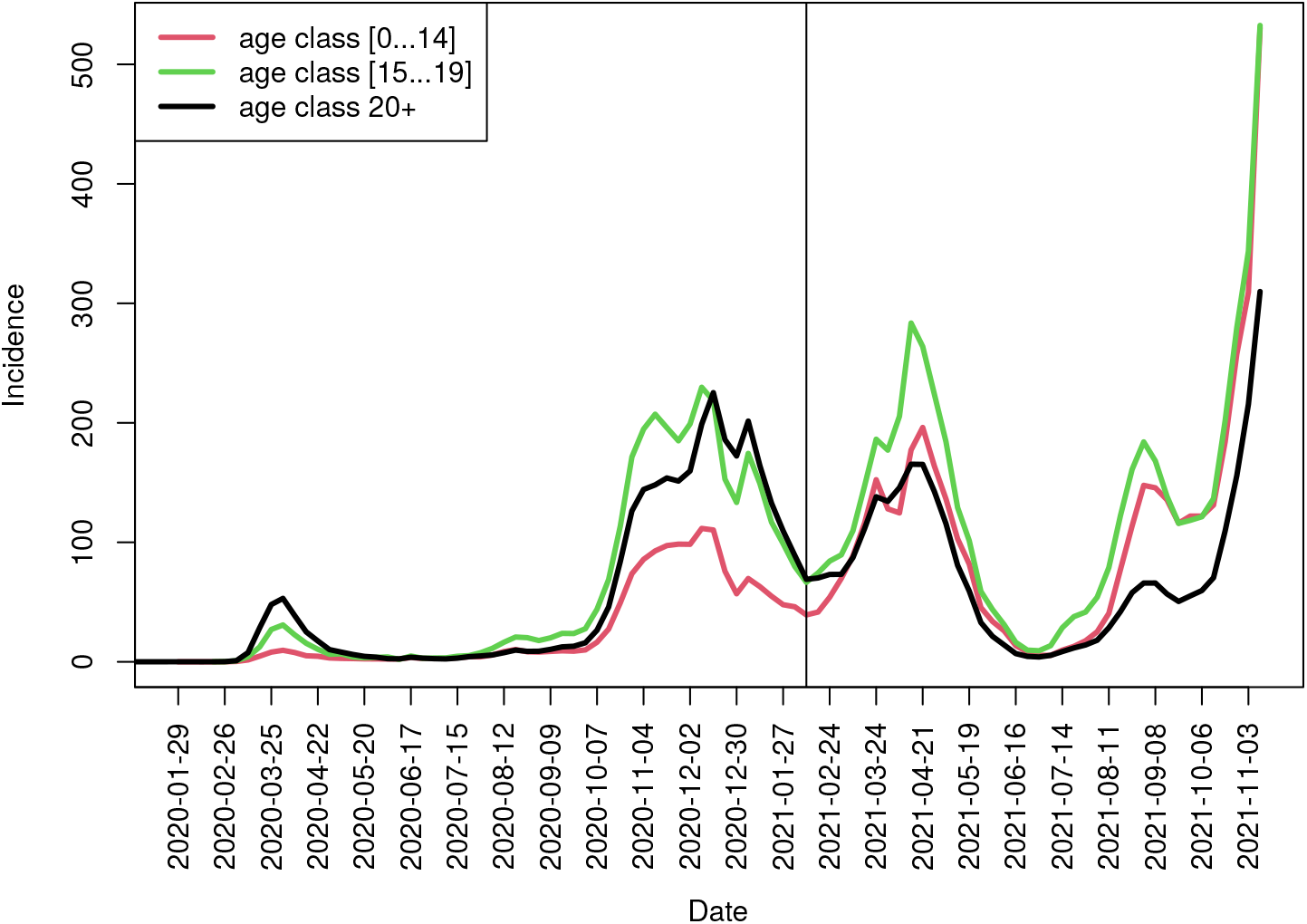
German-wide 7-day-incidence (per 100,000) time series for children, juveniles, and adults, respectively, sampled per week. Mutual cross-correlations are calculated for the two temporal intervals separated by the vertical line (at week 59).

**Figure 2:**
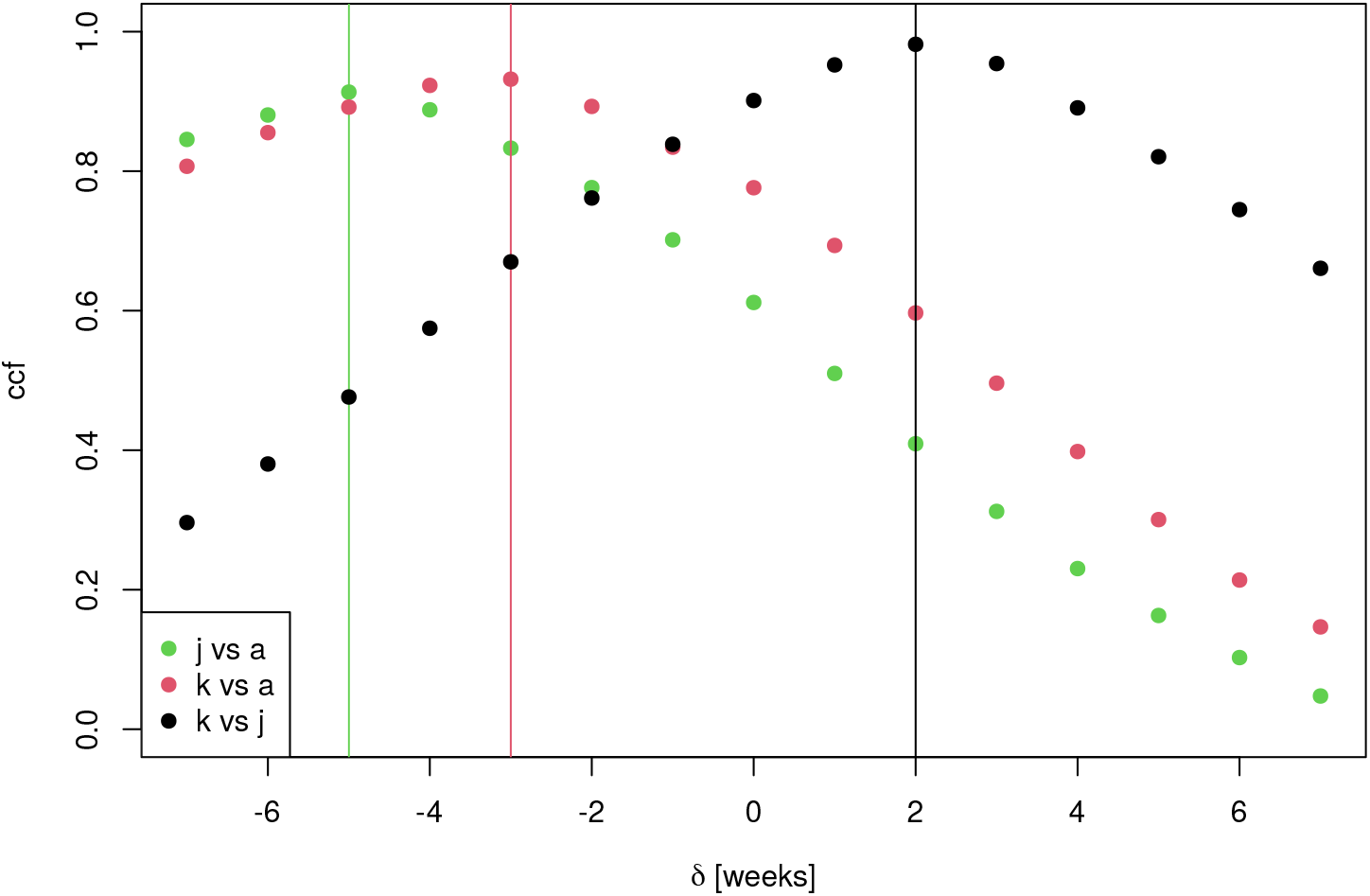
Cross-correlation functions by phase shift *δ* for *I*_*x*_(*t*+*δ*) versus *I*_*y*_(*t*) for the three (*x, y*)-pairs (*j, a*), (*k, a*), (*k, j*), respectively, during the first two waves (*t <* 60 weeks). For each pair, maximum *δ* is highlighted by a vertical line.

### 3.2 Cross-Correlations on the Local District Levels for the First Two Waves

Calculating the maximum cross-correlations between *I*_*j*_(*t* + *δ*) and *I*_*a*_(*t*) per district yields the distribution of phase shifts *δ* depicted in Table 1. It turns out, that 20% of the districts exhibit negative phase shifts (*δ <* 0) and 3% exhibit a positive phase shift of 1 week, which arguably is negligible. All districts which exhibit phase shifts *δ <* −1 weeks are listed in Table 2 along with the concrete values of *δ* as well as the corresponding maximum correlation coefficients. Furthermore, the 7-day incidence values at final observation time *t*_*f*_ are shown for each of the three sub-populations (age classes). Finally, the federate state to which the district belongs is listed in the last column.

**Table 1:** Distribution of phase shifts *δ* (in weeks) between the two incidence curves corresponding to the juvenile and adult sub-populations for the first two waves (*t <* 60 weeks) per district.

| $\delta$ | Freq |
| --- | --- |
| -6 | 1 |
| -5 | 3 |
| -4 | 3 |
| -3 | 3 |
| -2 | 13 |
| -1 | 61 |
| 0 | 315 |
| 1 | 12 |

**Table 2:** List of districts exhibiting phase shifts during the first two waves between *I*_*j*_ and *I*_*a*_ less than −1. Also shown are the phase shifts *δ* that maximize the correlation as well as the maximum correlation (corr). Further, the federate states to which the districts belong (last column) as well as the 7-day-incidence values per age class at the final observation time, *t*_*f*_ = 17 Nov 2021, are shown.

| District | $\delta$ | corr | $I_k(t_f)$ | $I_j(t_f)$ | $I_a(t_f)$ | Federate State |
| --- | --- | --- | --- | --- | --- | --- |
| SK Ansbach | -6 | 0.62 | 461.46 | 407.54 | 342.52 | Bayern |
| SK Aschaffenburg | -4 | 0.87 | 392.91 | 739.55 | 332.58 | Bayern |
| SK Memmingen | -2 | 0.74 | 648.93 | 1310.86 | 646.15 | Bayern |
| SK Berlin Lichtenberg | -2 | 0.92 | 414.52 | 269.67 | 166.42 | Berlin |
| LK Oberspreewald Lausitz | -3 | 0.91 | 2322.87 | 1288.53 | 595.20 | Brandenburg |
| SK Frankfurt/Oder | -2 | 0.89 | 788.78 | 394.74 | 288.17 | Brandenburg |
| LK Breisgau Hochschwarzwald | -4 | 0.71 | 474.53 | 305.30 | 262.40 | BW |
| LK Nordwestmecklenburg | -4 | 0.78 | 189.68 | 178.33 | 91.43 | MV |
| SK Rostock | -2 | 0.76 | 366.40 | 275.09 | 244.89 | MV |
| LK Friesland | -2 | 0.76 | 78.10 | 105.42 | 61.41 | Niedersachsen |
| SK Wilhelmshaven | -2 | 0.60 | 108.53 | 186.16 | 84.46 | Niedersachsen |
| LK Wesel | -2 | 0.93 | 321.82 | 141.12 | 140.64 | NRW |
| SK Mülheim/Ruhr | -2 | 0.88 | 220.59 | 186.17 | 108.70 | NRW |
| LK Bad Dürkheim | -3 | 0.85 | 308.84 | 133.67 | 181.14 | RP |
| SK Frankenthal | -2 | 0.77 | 540.85 | 398.58 | 190.04 | RP |
| LK Harz | -2 | 0.87 | 568.02 | 439.64 | 145.43 | SA |
| LK Saalekreis | -2 | 0.89 | 678.83 | 551.82 | 331.60 | SA |
| SK Dessau Roßlau | -2 | 0.78 | 738.09 | 1141.23 | 396.40 | SA |
| LK Ostholstein | -5 | 0.73 | 106.62 | 55.31 | 68.64 | SH |
| LK Plön | -5 | 0.63 | 112.38 | 79.83 | 66.87 | SH |
| LK Stormarn | -5 | 0.81 | 209.21 | 108.74 | 95.11 | SH |
| LK Eichsfeld | -3 | 0.86 | 434.33 | 744.25 | 412.30 | Thüringen |
| LK Sonneberg | -2 | 0.85 | 1021.05 | 1755.16 | 975.23 | Thüringen |

Analogously, the above procedure is applied to the maximum cross-correlations between *I*_*k*_(*t* + *δ*) and *I*_*a*_(*t*) (kids versus adults) which yields the results depicted in Table 3. A proportion of 19.2% of the districts exhibit negative phase shifts (*δ <* 0) and 6% exhibit a positive phase shift mostly of 1 week. As for the juveniles, the districts which exhibit phase shifts *δ <* −1 weeks are listed in Table 4 (with the same column structure as Table 2). Of note, there are considerably more districts with phase shifts *δ <* −1 weeks for kids (44) compared to juveniles (23).

**Table 3:** Distribution of phase shifts *δ* (in weeks) between the two incidence curves corresponding to the children and adult sub-populations for the first two waves (*t <* 60 weeks) per district.

| $\delta$ | Freq |
| --- | --- |
| -6 | 4 |
| -5 | 5 |
| -4 | 3 |
| -3 | 14 |
| -2 | 18 |
| -1 | 35 |
| 0 | 307 |
| 1 | 23 |
| 2 | 1 |
| 4 | 1 |

**Table 4:** List of districts exhibiting phase shifts during the first two waves between *I*_*k*_ and *I*_*a*_ less than −1. Specification of columns analogous to Table 2.

| LK | $\delta$ | corr | $I_k$ | $I_j$ | $I_a$ | BL |
| --- | --- | --- | --- | --- | --- | --- |
| LK Bad Kissingen | -2 | 0.83 | 948.88 | 670.44 | 424.25 | Bayern |
| LK Bayreuth | -2 | 0.81 | 571.30 | 553.51 | 420.44 | Bayern |
| LK Dingolfing Landau | -2 | 0.71 | 1267.41 | 1412.07 | 959.51 | Bayern |
| LK Rhön Grabfeld | -3 | 0.79 | 868.35 | 736.79 | 388.51 | Bayern |
| LK Straubing Bogen | -2 | 0.79 | 1325.91 | 945.07 | 656.44 | Bayern |
| LK Wunsiedel/Fichtelgebirge | -4 | 0.57 | 637.61 | 538.83 | 448.56 | Bayern |
| SK Ansbach | -5 | 0.67 | 461.46 | 407.54 | 342.52 | Bayern |
| SK Aschaffenburg | -2 | 0.84 | 392.91 | 739.55 | 332.58 | Bayern |
| SK Hof | -2 | 0.85 | 399.73 | 735.65 | 274.71 | Bayern |
| SK Kaufbeuren | -3 | 0.74 | 583.28 | 689.66 | 429.57 | Bayern |
| SK Kempten | -3 | 0.76 | 1057.73 | 1035.26 | 683.42 | Bayern |
| SK Rosenheim | -2 | 0.80 | 629.36 | 954.65 | 552.78 | Bayern |
| SK Berlin Reinickendorf | -2 | 0.89 | 687.90 | 350.37 | 278.65 | Berlin |
| LK Oberhavel | -3 | 0.85 | 567.51 | 266.18 | 190.43 | Brandenburg |
| LK Spree Neiße | -5 | 0.79 | 1365.36 | 745.91 | 477.10 | Brandenburg |
| SK Frankfurt/Oder | -3 | 0.76 | 788.78 | 394.74 | 288.17 | Brandenburg |
| LK Rheingau Taunus Kreis | -2 | 0.87 | 238.67 | 98.19 | 136.49 | Hessen |
| LK Wetteraukreis | -3 | 0.88 | 255.28 | 225.57 | 171.44 | Hessen |
| LK Nordwestmecklenburg | -3 | 0.83 | 189.68 | 178.33 | 91.43 | MV |
| LK Werra Meißner Kreis | -2 | 0.78 | 352.49 | 155.69 | 218.35 | MV |
| LK Friesland | -2 | 0.78 | 78.10 | 105.42 | 61.41 | Niedersachsen |
| LK Goslar | -3 | 0.71 | 154.69 | 170.39 | 94.78 | Niedersachsen |
| LK Hameln Pyrmont | -2 | 0.89 | 265.16 | 112.69 | 75.49 | Niedersachsen |
| LK Oldenburg | -6 | 0.65 | 141.88 | 71.69 | 138.46 | Niedersachsen |
| LK Wesermarsch | -6 | 0.57 | 190.11 | 87.95 | 102.20 | Niedersachsen |
| SK Neustadt/Weinstraße | -2 | 0.78 | 436.44 | 285.25 | 192.00 | RP |
| LK Börde | -4 | 0.83 | 502.03 | 656.36 | 232.62 | SA |
| LK Harz | -2 | 0.85 | 568.02 | 439.64 | 145.43 | SA |
| LK Saalekreis | -3 | 0.90 | 678.83 | 551.82 | 331.60 | SA |
| SK Dessau Roßlau | -2 | 0.77 | 738.09 | 1141.23 | 396.40 | SA |
| LK Leipzig | -3 | 0.91 | 1982.07 | 1507.82 | 754.21 | Sachsen |
| LK Nordsachsen | -3 | 0.88 | 1144.50 | 1275.98 | 542.90 | Sachsen |
| SK Chemnitz | -2 | 0.96 | 1015.10 | 919.23 | 554.69 | Sachsen |
| SK Leipzig | -3 | 0.90 | 781.64 | 533.32 | 399.69 | Sachsen |
| LK Herzogtum Lauenburg | -6 | 0.75 | 233.64 | 130.12 | 113.41 | SH |
| LK Ostholstein | -5 | 0.73 | 106.62 | 55.31 | 68.64 | SH |
| LK Pinneberg | -6 | 0.76 | 181.63 | 136.26 | 79.91 | SH |
| LK Plön | -5 | 0.66 | 112.38 | 79.83 | 66.87 | SH |
| LK Gotha | -4 | 0.71 | 889.78 | 1188.40 | 740.43 | Thüringen |
| LK Greiz | -2 | 0.72 | 554.98 | 797.66 | 498.92 | Thüringen |
| LK Kyffhäuserkreis | -5 | 0.68 | 599.82 | 873.95 | 554.20 | Thüringen |
| LK Schmalkalden Meiningen | -3 | 0.87 | 807.82 | 1137.54 | 642.92 | Thüringen |
| LK Sonneberg | -3 | 0.81 | 1021.05 | 1755.16 | 975.23 | Thüringen |
| LK Wartburgkreis | -2 | 0.90 | 581.96 | 963.16 | 489.67 | Thüringen |

Apparently, there is no clear pattern observable in Tables 2 and 4. At a first glance, it looks like a random mixture of federate states and current epidemic activities reflected by the incidence values at final observation time, which will be analyzed in more detail below. However, we are currently unaware of the local policies of these districts which would likely be informative. Main finding so far is that within 20% of the districts as well as for the German-wide average, the epidemic activities of both the juvenile as well as the children sub-populations reveal similar shapes as the activities of the adult sub-populations, however, the incidence time series exhibit negative phase shifts: The juveniles’ and the children’s epidemic activities are ahead of the adults’ activity. Of note, this result holds for observed and registered cases only. The amount of unobserved cases most likely depends on the age class. However, the results are robust contingent on the assumption that the proportion of unobserved cases remains approximately constant over the observation time.

### 3.3 Prediction of Current Epidemic Activity by Age-Specific Phase Shifts

A linear regression modeling the current (*t*_*f*_ =final observation time 17 Nov 2021) local 7-day-incidences of the total (*I*_*total*_(*t*_*f*_)) or, alternatively, the adult (*I*_*a*_(*t*_*f*_)) sub-population dependent on the phase shifts observed during the first two waves yields the results depicted in Figure 3. Specifically, the phase shift between the incidence curves corresponding to the youngest and the adult sub-populations predicts the current incidence *I*_*total*_(*t*_*f*_, *δ*) ∝ *δ* with slope 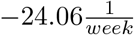 and p-value *p* = 0.09. In other words, observing an epidemic activity of children ahead of the corresponding activity of the adults during the first two waves moderately predicts an incidence above the average during the fourth wave. The inverse holds for the juveniles but below significance (slope 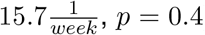). Interestingly, predicting the current incidence of the adults instead of the total incidence by the shift between the curves corresponding to kids and adults during the first half of the epidemic, yields slope 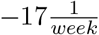 and slightly smaller p-value 0.077. Again, the negative phase shift of the curve corresponding to the juvenile population does not contribute to the prediction of the current activity of the adult sub-population (slope 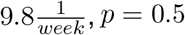).

**Figure 3:**
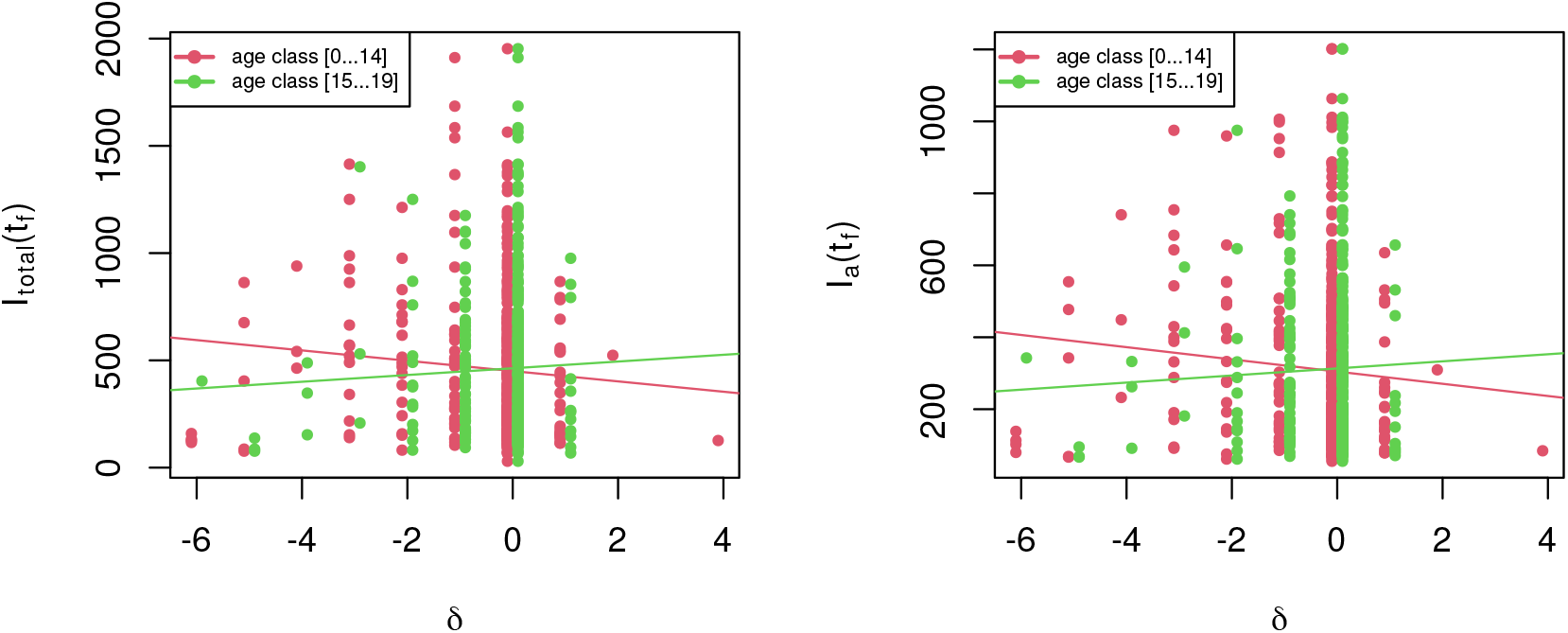
Linear regression predicting the current (*t*_*f*_ = 17 Nov 2021) total incidence (left panel) and the incidence of the adult sub-population (right panel), respectively, observed in the 411 German districts by the phase shift between *I*_*j*_(*t*) and *I*_*a*_(*t*) (green) or alternatively by the phase shift between *I*_*k*_(*t*) and *I*_*a*_(*t*) (red) for *t <* 60 weeks.

We so far conclude, an epidemic activity of children ahead of the adult activity during the first epidemic phase considerably predicts a higher overall activity during a later epidemic phase and even more significantly a higher activity of the adult sub-population.

### 3.4 Cross-correlations after the second wave

On the global (German-wide) level, the three mutual cross-correlations between the three age-specific incidence time series calculated for the second half of the epidemic (*t >* 59 weeks) yield correlation coefficients as a function of phase *δ* as depicted in Figure 4. A maximum correlation close to 1 is obtained for phase shift *δ* = 0 for all three pairs of incidence curves.

**Figure 4:**
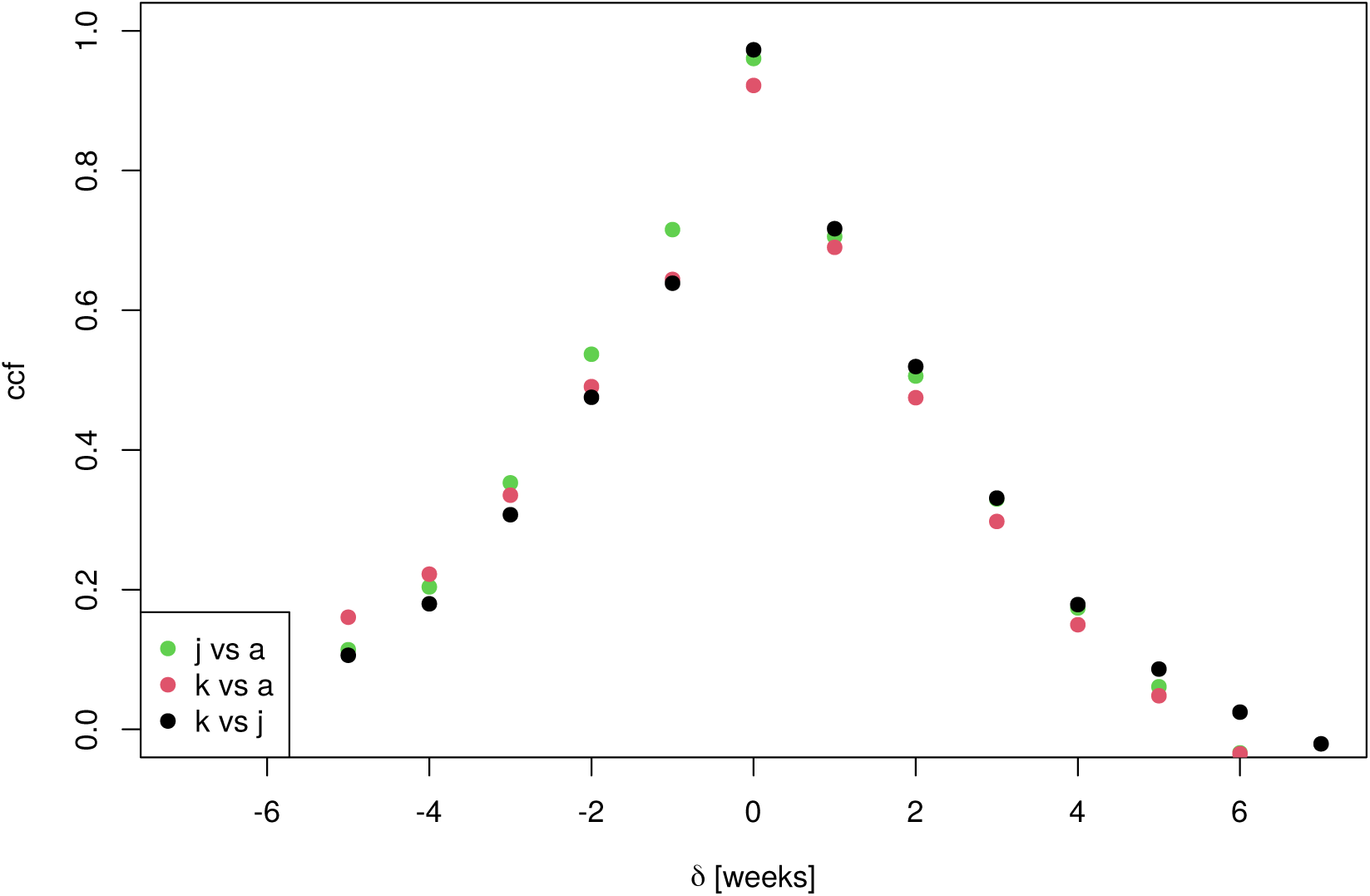
Cross-correlation functions by phase shift *δ* for *I*_*x*_(*t*+*δ*) versus *I*_*y*_(*t*) for the three (*x, y*)-pairs (*j, a*), (*k, a*), (*k, j*), respectively, during the second half of the epidemic (*t >* 59 weeks).

Thus, the dynamic patterns are almost perfectly synchronous with an apparently very similar shape, although the amplitudes differ substantially. Obviously, both containment strategies as well as testing frequencies unfold an almost equal impact on sub-populations of all age classes.

Likewise on the district level, phase shifts during the second half the epidemic differing from *δ* = 0 are rare, as can be seen in Tables 5 and 6. We conclude that the activity patterns corresponding to different age classes are more or less synchronous roughly from week *t* = 60 onward.

**Table 5:** Distribution of phase shifts *δ* (in weeks) between the two incidence curves corresponding to the juveniles and adult sub-populations for the second half of the epidemic (*t >* 59 weeks) per district.

| $\delta$ | Freq |
| --- | --- |
| -2 | 1 |
| -1 | 7 |
| 0 | 400 |
| 1 | 3 |

**Table 6:** Distribution of phase shifts *δ* (in weeks) between the two incidence curves corresponding to the children and adult sub-populations for the second half of the epidemic (*t >* 59 weeks) per district.

| $\delta$ | Freq |
| --- | --- |
| -3 | 2 |
| -1 | 6 |
| 0 | 402 |
| 1 | 1 |

## 4 Discussion

In Germany, children and juveniles unfold a SARS-CoV-2 epidemic activity which is ahead of the adult activity during the first phase of the epidemic (comprising two waves) by means of maximizing the time-delayed cross-correlation between the corresponding incidence time series. Furthermore, the phase shift between the children’s incidence curve and the adults’ curve has predictive power for both the epidemic overall as well as adults’ activity in a later epidemic period. Interpretations have to be given with utmost care. The analysis is based on registered (observed) incidence only which may differ in an age-dependent way from the “true” incidence. Furthermore, the ratio of unobserved to observed cases per week may be subject to temporal changes depending on current policies with respect to test frequencies. However, assuming that children are under-tested due to lack of symptoms may render the results even as underestimated.

Having said that, with due caution, it appears that children and juveniles are “drivers” of the epidemic at least during the first pandemic year. Vaccination coverage of the adult sub-population may be a crucial factor behind the substantially reduced phase shifts between the age-dependent incidence curves during the second half of the epidemic. After the second wave, the epidemic activities of different age classes appear to be much more synchronized.

It should be mentioned that policies regarding opening/closure of schools and testing procedures have to be based on much more factors as, for example, learning loss and sociopsychological effects. Children have a low risk of severe disease progression which has to be balanced by the risk of severe psychological harm through isolation. Nevertheless, for future decision making, the thus far underestimated role of kids in driving the epidemic should urgently find more attention and it demands for better protective measures and an increased frequency of more sensitive testing. The districts exhibiting negative phase shifts of the children’s incidence curve are particularly addressed to investigate the underlying causes in more detail to eventually inform and improve the respective decision making.

## Data Availability

Publicly available data provided by survstat.rki.de have been used exclusively.

## Data

Publicly available data provided by [9] have been used exclusively.

## Abbreviations

BW: Baden Wü rttemberg
MV: Mecklenburg-Vorpommern
NRW: Nord-Rhein-Westfalen
RP: Rheinland-Pfalz
SA: Sachsen-Anhalt
SH: Schleswig-Holstein

